# ProbeFit: one-seizure qualification and personalisation of multimodal wearable seizure detectors

**DOI:** 10.64898/2026.11.17.26368305

**Authors:** Han Cui, Yuge Zhu, Shaojia Huang, Tao Xu

**Affiliations:** Department of Biomedical Engineering, College of Engineering, Shantou University, 243 Daxue Road, Shantou 515063, China; School of Intelligent Manufacturing and Aeronautics, Zhuhai College of Science and Technology, 8 Anji East Road, Jinwan District, Zhuhai 519041, China

**Keywords:** epilepsy, wearable, seizure detection, personalisation, behind-the-ear EEG, patient selection

## Abstract

**Background and Objective:** Missed seizures hinder assessment of seizure frequency and treat-ment. Although wearables can detect seizures automatically, performance varies between pa-tients. Before deployment, it remains difficult to identify suitable patients, and personalisation usually requires several recorded seizures. We propose ProbeFit to predict later seizure sensi-tivity and personalise a wearable detector from the first recorded seizure.

**Methods:** We used multicentre SeizeIT2 recordings of behind-the-ear EEG, ECG, EMG and movement from 125 patients. Two generic detectors’ peak responses to the first seizure gener-ated a qualification probability. The same seizure and preceding background personalised the detector. ProbeFit was developed on 96 patients and applied unchanged to 29 held-out patients. Expert-labelled video-EEG seizures provided the reference. Evaluation compared qualification with the first-seizure heart-rate rule and measured discrimination, calibration and net benefit, and it compared later sensitivity of the generic and personalised detectors at equal false-alarm rates.

**Results:** Among 27 held-out patients with 152 later seizures, qualification correlated with later sensitivity at Spearman ρ = 0.38, with one-sided p = 0.033. It identified patients with later sensitivity ≥ 0.5 with an area under the ROC curve (AUC) of 0.74. Across all 97 evaluable patients, AUC was 0.76 against 0.59 for the heart-rate rule, a difference of 0.17 with a 95% confidence interval of 0.05 to 0.29. At the qualification threshold of 0.5, ProbeFit withheld deployment from 9 of 15 held-out patients below the sensitivity criterion, and its net benefit was 0.074 against −0.111 for selecting everyone. At equal false-alarm rates between 0.25 and 2 per day, personalisation raised later sensitivity by 2.2 percentage points in held-out patients and by 6.3 points in development patients. Both confidence intervals excluded zero, and no held-out patient lost sensitivity.

**Conclusions:** ProbeFit uses the first recorded seizure to predict whether a wearable detector will detect at least half of a patient’s later seizures, and it personalises that detector without waiting for further seizures. It supports patient selection and adaptation during the first moni-toring admission.

## 1. Introduction

Epilepsy affects more than 45 million people worldwide [1]. Its recurrent seizures vary in onset and clinical presentation [2, 3], and treatment decisions depend on documenting them reliably [4, 5]. Video-EEG provides the reference standard, while wearables support prolonged monitoring outside hospital [5, 6]. Patient diaries miss many seizures, particularly those that impair awareness or occur during sleep [7, 8]. Unnoticed seizures also complicate safety mon-itoring, including the recognition of nocturnal tonic-clonic seizures associated with sudden un-expected death in epilepsy [9].

Wearable detectors combine movement and autonomic signals [4, 10], while behind-the-ear EEG (bte-EEG) and electrocardiography (ECG) capture electrical and cardiac changes during focal seizures [11–14]. SeizeIT2 provides multicentre recordings of these signals from 125 pa-tients [15]. Generic detectors use feature-based classifiers or neural networks [10–12, 16, 17], with event-based evaluation supported by SzCORE [18]. Patient-specific methods learn seizure signatures [19–21] or adjust thresholds to individual recordings [22–24]. Patient selection offers another approach: pronounced ictal heart-rate changes identify patients suited to heart-rate-var-iability detectors [25, 26]. A prospective study applied this rule to the first recorded seizure [27], followed by its evaluation in SeizeIT2 [28]. However, heart-rate change does not identify every patient whose seizures a wearable detector can detect [29].

These approaches leave a practical question: what can the first recorded seizure tell us about the detector a patient will wear? In SeizeIT2, detection depended on the ictal EEG pattern and accompanying tachycardia [30]. Thus, selection for a multimodal detector should account for its response to both signals. Methods that assess uncertainty in individual EEG predictions ad-dress a different decision, namely which segments require clinical review [31]. Personalisation must also respect the deployment time: training data available afterwards cannot guide a deci-sion made at the first seizure.

We therefore propose ProbeFit, which uses the generic detectors’ responses to the first rec-orded seizure to predict whether a personalised detector will detect at least half of the patient’s later seizures. The same seizure and preceding background recording personalise the detector. We evaluate its discrimination, calibration and deployment net benefit in held-out SeizeIT2 patients against the published first-seizure heart-rate rule, test whether its prediction persists for temporally distant seizures and compare personalised and generic detection at equal false-alarm rates.

## 2. Methods

### 2.1. Problem formulation and overview

The first recorded seizure is the index seizure; later seizures begin at least 60 s after its offset. For n later seizures with h detections, sensitivity is S = h/n. A good-detection patient has S ≥ 0.5. Usable detection additionally requires FP ≤ max(1, H/24), where FP denotes false alarms over H evaluation hours.

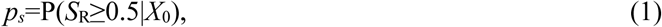

Here *S*_R_ is the later sensitivity of the index-personalised detector, and *X*_0_ contains the record-ing available for qualification. ProbeFit combines a Detector-Response Probe, Index-Anchored Refitting and a Qualification Gate, as illustrated in Fig. 1. The probe converts two generic de-tectors’ index-seizure responses into *p_s_*. Refitting adapts the gradient-boosted detector using that seizure and preceding background. The gate selects patients with *p_s_* ≥ 0.5. Table 1 defines the notation.

**Fig. 1.**
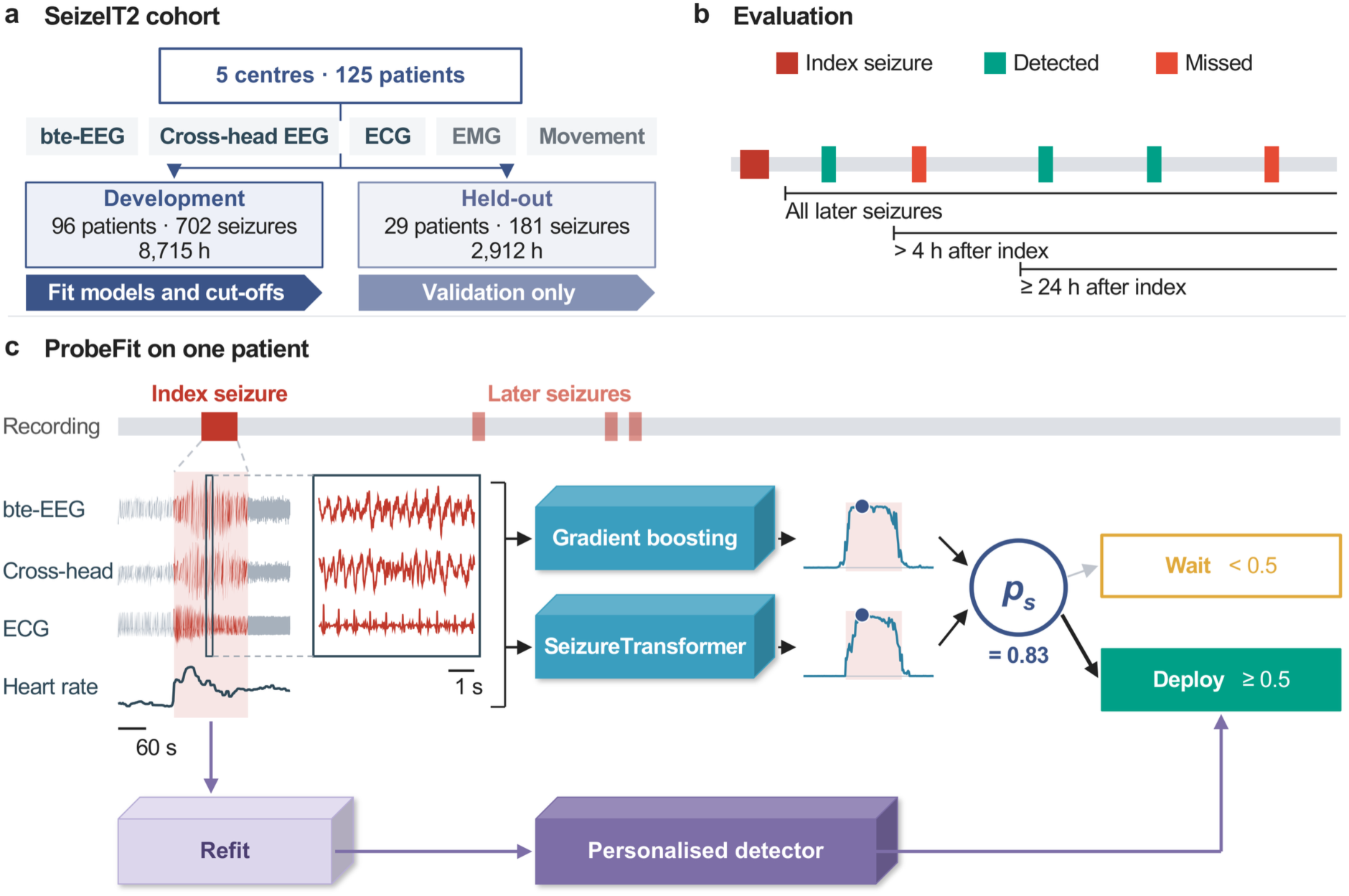
One-seizure qualification and personalisation with ProbeFit. (a) Cohort, signals and patient split. (b) Index and later seizures; good detection means later sensitivity ≥ 0.5. (c) Example recording and enlarged index seizure, marked in red. Dots show generic detectors’ peak responses; their combination gives *p_s_*, with deployment selected at *p_s_* ≥ 0.5. Refitting uses the index seizure, preceding background and training patients.

**Table 1.** Symbols used in the paper.

| **Symbol** | **Meaning** |
|---|---|
| H | Hours of evaluation |
| n | Number of later seizures |
| h | Number of detected later seizures |
| FP | Number of false alarms |
| *m<sub>k</sub>* | Peak smoothed score of detector k from 30 s before to 60 s after the index seizure |
| *l<sub>k</sub>* | Clipped logit of *m<sub>k</sub>* |
| *z<sub>k</sub>* | Standardised logit |
| *p<sub>s</sub>* | Qualification probability |
| *τ*<sub>G</sub>, *τ*<sub>R</sub> | Thresholds of the generic and index-personalised detectors |
| S, *S*<sub>R</sub> | Later sensitivity; that of the detector the patient will wear |
| *X*<sub>0</sub> | Recording available for qualification, including the scoring margin |
| D, I, B | Training sample, index-seizure windows and pre-index background windows |
| *w<sub>i</sub>*, λ, W<sup>+</sup> | Window weights in Eq. (4) |
| ρ | Spearman correlation |

### 2.2. Generic detectors and event scoring

We used a histogram-based gradient-boosting classifier [32] and SeizureTransformer [16]. The former used spectral and temporal bte-EEG features, ECG-derived heart-rate features, EMG and movement features, with local temporal context. It had 300 iterations, learning rate 0.08 and 31 leaves, and balanced class weights. SeizureTransformer was retrained from random initialisation on two EEG channels and ECG, using 60-s windows at 256 Hz. Supplementary Methods S1 gives preprocessing, feature extraction and training details.

Five-fold cross-validation grouped by patient generated development scores from models trained on other patients. Held-out patients received one fold model through a fixed balanced mapping. Scores were averaged into 2-s windows and smoothed with a centred five-window mean. Above-threshold windows formed events; gaps shorter than 90 s were merged and events longer than 5 min split. A detection overlapped the interval from 30 s before seizure onset to 60 s after offset; other events were false alarms. Generic and personalised thresholds were the lowest giving one pooled false alarm per day in development evaluation periods. Development thresholds were cross-fitted using the other four folds, whereas held-out thresholds used all development patients.

### 2.3 Detector-Response Probe

Each detector contributed its maximum smoothed score *m_k_* from 30 s before index onset to 60 s after offset. We clipped scores before taking logits, with ε = 10^−6^ for gradient boosting and 10^−4^ for SeizureTransformer:

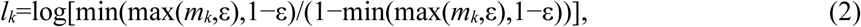

Development means and standard deviations converted these logits into *z*_1_ and *z*_2_. Logistic regression combined them:

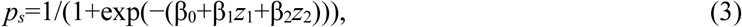

The outcome was later sensitivity ≥ 0.5 for the index-personalised detector. The model used L2 regularisation with C = 1. Development probabilities came from leave-one-patient-out fit-ting among the 70 evaluable patients, whereas held-out probabilities used the fixed develop-ment model.

### 2.4 Index-Anchored Refitting and Qualification Gate

We refitted the gradient-boosted detector on its original training sample D, index-seizure windows I and up to 20,000 background windows B ending 30 s before index onset (median 9.8 h in development):

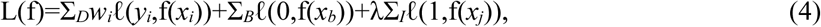

Here ℓ is logistic loss, *w_i_* balances classes in D, and λ = 0.1 W⁺/|I|, where W⁺ is the total positive weight in D. Thus, the index seizure contributes 10% of the original positive weight. Background windows adapt the detector to the patient’s non-seizure signals. The gate selects the personalised detector when

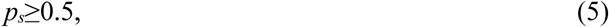

which assigns equal weight to correct and incorrect selection in the net-benefit analysis. Sup-plementary Methods S2 provides the algorithm.

### 2.5. Data and comparator

We used SeizeIT2 version 1.1.0, OpenNeuro ds005873, recorded during video-EEG moni-toring in five European centres [15]. Sensor Dot recorded bte-EEG, cross-head EEG, ECG, EMG, acceleration and rotation. For the sixteen patients without a cross-head channel, we used their second device EEG channel instead. Neurologists annotated seizures on simultaneous video-EEG. Following the split of the release [15], sub-001–096 formed the development set and sub-097–125 the held-out set. Later-seizure evaluation required at least one seizure after the index event.

The comparator was the first-seizure heart-rate rule [25, 27, 28]. R peaks were obtained with a band-pass and squared-derivative detector [33]. We removed RR intervals outside 0.3–2.0 s or differing by more than 20% from the surrounding five-interval median. The rule was positive for heart-rate changes exceeding 50 beats/min across the index seizure’s RR intervals, extended symmetrically to 100 intervals when necessary. Unusable ECG yielded a negative decision.

### 2.6 Evaluation and statistical analysis

Held-out patients were analysed once, after all models and thresholds had been fixed. The rank correlation between *p_s_* and later sensitivity was tested one-sided with 10,000 permutations within fold strata. AUC and its difference from the comparator were reported with patient-level bootstrap confidence intervals using 5,000 resamples. We calculated deployment net benefit at probability threshold 0.5 [34] and assessed calibration by mean predicted probability, observed outcome frequency and calibration slope [35], with bootstrap intervals. A pooled analysis of all 97 evaluable patients combined out-of-sample development and held-out probabilities and compared ProbeFit with the index seizure’s annotated duration and type.

We repeated evaluation for seizures more than 4 h or at least 24 h after the index seizure. A seizure-free control used the gradient-boosted detector’s median peak over 20 random 2-min spans at least 1 h from seizures in development patients. To compare detectors at equal false-alarm rates, each threshold was the lowest value giving a target pooled rate, reset in each of 1,000 bootstrap resamples, and sensitivity differences were averaged over 0.25–2 false alarms/day. Reporting follows TRIPOD+AI where applicable [36]. Analyses used Python 3.11.18, scikit-learn 1.9.1, PyTorch 2.15.0 and MNE-Python 1.13.2.

## 3. Results

### 3.1. Cohort and generic detection

The development and held-out sets contained 96 and 29 patients, respectively. Their record-ings totalled 8,715 and 2,912 h, with 702 and 181 seizures. Later-seizure evaluation included 70 development patients with 605 seizures and 27 held-out patients with 152 seizures. Table 2 summarises the cohort.

**Table 2.** Patients, recordings and seizures.

|  | **Development** | **Held-out** |
|---|---|---|
| Patients | 96 | 29 |
| Female / male | 51 / 45 | 11 / 18 |
| Recording hours | 8,715 | 2,912 |
| Hours per patient, median (range) | 90 (1–397) | 99 (3–277) |
| Seizures | 702 | 181 |
| Seizures per patient, median (range) | 3 (1–75) | 4 (1–37) |
| Focal impaired awareness | 289 (41%) | 102 (56%) |
| Focal aware | 274 (39%) | 42 (23%) |
| Focal, awareness unknown | 105 (15%) | 10 (6%) |
| Focal to bilateral tonic-clonic | 33 (5%) | 22 (12%) |
| Unknown onset | 1 (<1%) | 5 (3%) |
| Seizures during sleep | 274 (39%) | 93 (51%) |
| Patients with temporal-lobe predominant onset | 57 (59%) | 18 (62%) |
| Evaluated patients | 70 | 27 |
| Later seizures | 605 | 152 |
| Hours of evaluation | 4,578 | 1,904 |

Seizure types are those annotated in the release. Predominant onset is the most frequent onset region of a patient’s seizures; temporal-lobe onset includes fronto-temporal and other regions involving the temporal lobe. Evaluated patients had an index seizure followed by at least one later seizure. The public release does not give age or centre per patient.

Generic detection varied between patients. In development, pooled later sensitivity was 21.5% at 2.0 false alarms/day. Half the evaluable patients had no later seizures detected, whereas 24 had at least half detected. Detection met both sensitivity and false-alarm criteria in 27% of patients, as shown in Fig. 2a. Detection reached 94% for focal to bilateral tonic-clonic seizures but 11% for focal non-motor seizures, as reported in Supplementary Table S1.

**Fig. 2.**
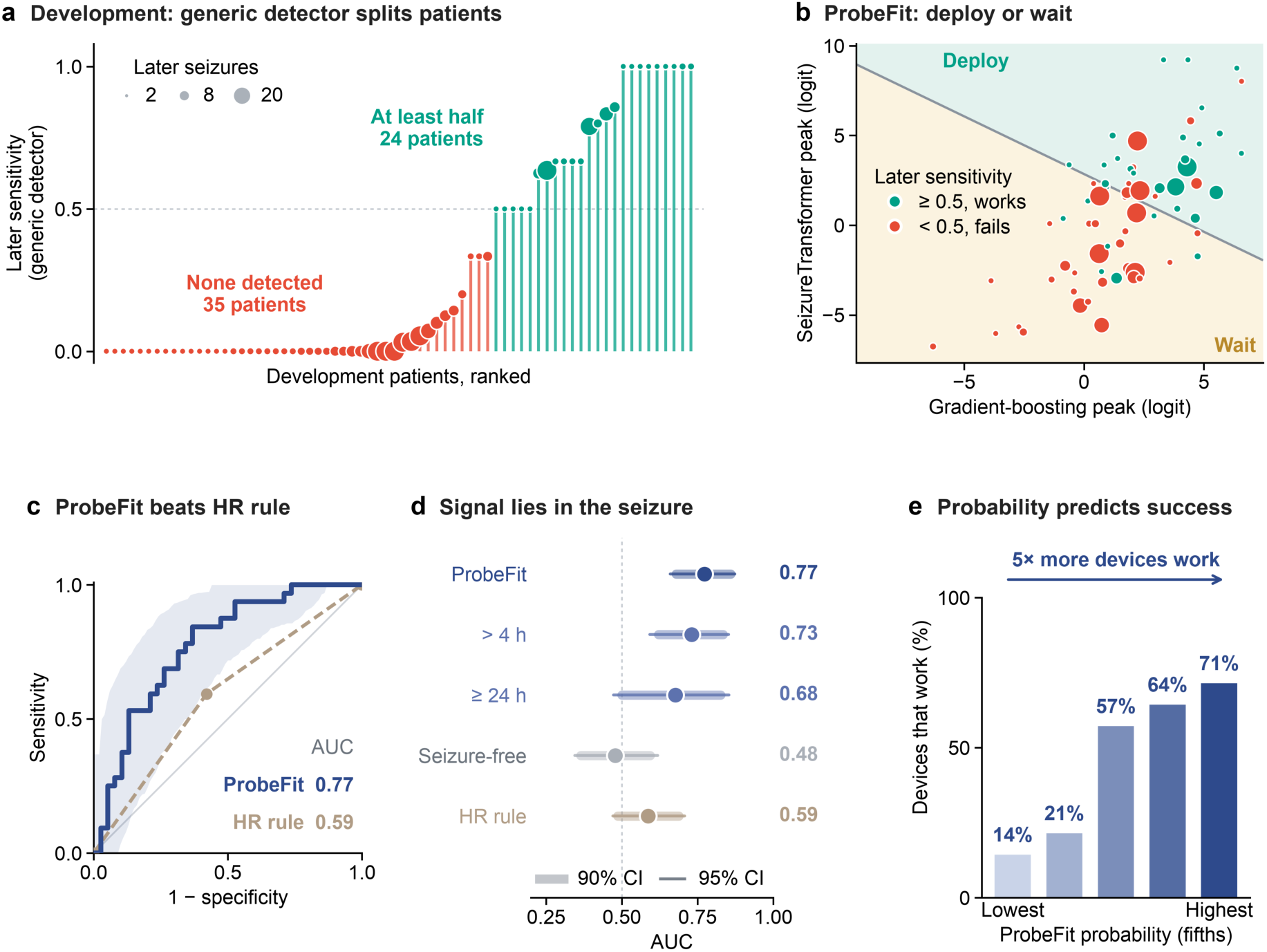
Index-seizure responses predict later detection in development patients. (a) Generic sensitivity per patient; dot size represents later-seizure count and green indicates sensitivity ≥ 0.5. (b) Detector peaks, coloured by per-sonalised later sensitivity; the line marks *p_s_* = 0.5. (c) ROC curves with ProbeFit’s 95% bootstrap band and the heart-rate rule’s operating point. (d) AUC for temporal restrictions, seizure-free control and heart-rate rule; thick and thin bars show 90% and 95% bootstrap intervals. (e) Good-detection proportions in five groups ordered by *p_s_*.

### 3.2. Qualification in development patients

The out-of-sample probability identified good-detection patients with AUC 0.77, compared with 0.59 for the heart-rate rule. Individual detector peaks also discriminated these patients, as shown in Fig. 2b–c. Good-detection patients comprised 67% of those selected by ProbeFit and 27% of those not selected. Their proportion increased from 14% in the lowest probability group to 71% in the highest group in Fig. 2e.

The prediction persisted beyond nearby seizures: AUC was 0.73 among 62 patients with sei-zures more than 4 h after the index event. In contrast, seizure-free peaks gave AUC 0.48, as shown in Fig. 2d.

### 3.3. Held-out validation

Applied unchanged to held-out patients, *p_s_* correlated with later sensitivity at ρ = 0.38, with one-sided permutation p = 0.033. AUC was 0.74, with a 95% confidence interval of 0.52–0.91, as shown in Fig. 3a–b. The AUC difference from the heart-rate rule was 0.15, with an interval of −0.10 to 0.38. For seizures more than 4 h and at least 24 h after the index event, AUC was 0.70 in both analyses in Fig. 3c. Across all 97 evaluable patients, ProbeFit reached AUC 0.76 against 0.59 for the heart-rate rule, a difference of 0.17 with an interval of 0.05–0.29. The probe added information to the index seizure’s duration and type, with likelihood-ratio p = 0.001, but not the reverse, with p = 0.55, as reported in Supplementary Tables S2 and S3.

**Fig. 3.**
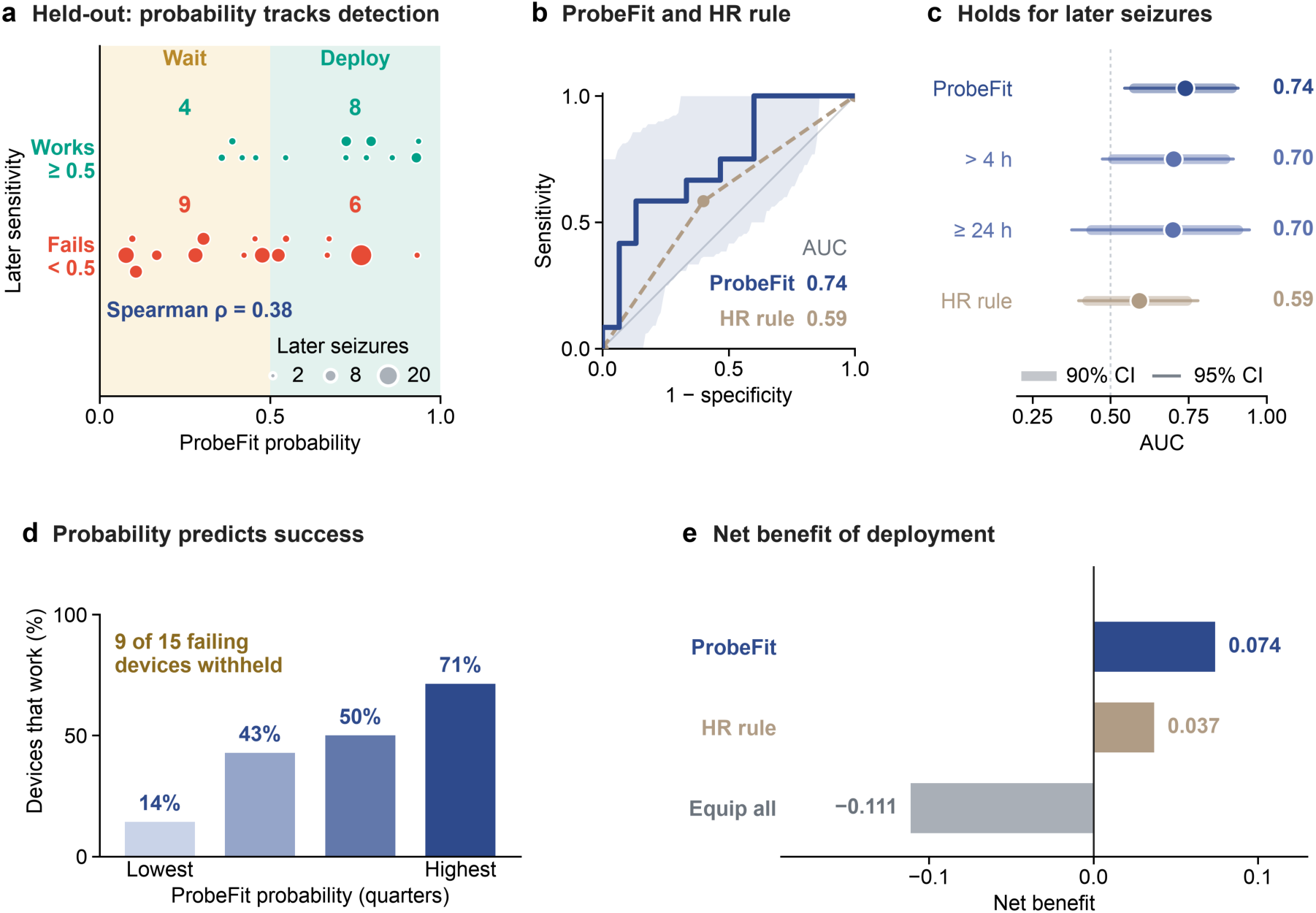
Qualification in held-out patients. (a) Probabilities grouped by personalised sensitivity ≥ 0.5 (Works) or < 0.5 (Fails); dot size represents later-seizure count. Shading marks selection regions; ρ relates probability to sensi-tivity. (b) ROC curves. (c) AUC under temporal restrictions, with intervals as in Fig. 2d. (d) Good-detection pro-portions in four groups ordered by *p_s_*. (e) Net benefit at probability threshold 0.5 for ProbeFit, the heart-rate rule and selecting everyone.

At the fixed gate, 57% of selected patients were good-detection patients, compared with 31% of those not selected. ProbeFit selected 8 of 12 good-detection patients and withheld deploy-ment from 9 of 15 patients below the sensitivity criterion. Its net benefit was 0.074, compared with 0.037 for the heart-rate rule and −0.111 for selecting everyone, as shown in Fig. 3d–e. Mean predicted probability was 0.53 against an observed proportion of 0.44; Table 3 reports calibration and performance measures.

**Table 3.** Main measures in development and held-out patients.

| **Measure** | **Development** | **Held-out** |
|---|---|---|
| AUC of *p<sub>s</sub>* | 0.77 (0.66 to 0.88) | 0.74 (0.52 to 0.91) |
| AUC of single-detector peaks, gradient-boosted / Sei-zureTransformer | 0.73 / 0.80 | 0.75 / 0.74 |
| AUC of the heart-rate rule | 0.59 (0.47 to 0.70) | 0.59 (0.41 to 0.78) |
| Spearman ρ, *p<sub>s</sub>* vs later sensitivity | 0.49 (0.28 to 0.67) | 0.38 (0.03 to 0.67) |
| Good-detection patients among deployed | 67% | 57% |
| Good-detection patients among not deployed | 27% | 31% |
| Net benefit, deployment by *p<sub>s</sub>* | 0.157 (0.000 to 0.314) | 0.074 (−0.222 to 0.333) |
| Net benefit, heart-rate rule | 0.043 | 0.037 |
| Net benefit, deployment to all | −0.086 | −0.111 |
| Mean *p<sub>s</sub>* / observed proportion | 0.46 / 0.46 | 0.53 / 0.44 |
| Calibration slope | 0.87 (0.40 to 1.75) | 0.77 (0.17 to 2.11) |
| Pooled later sensitivity, generic → personalised | 21.5% → 30.1% | 13.8% → 19.1% |
| False alarms per day, generic → personalised | 2.0 → 1.5 | 0.39 → 0.77 |
| Sensitivity gain at equal false alarms, 0.25–2/day (points) | 6.3 (1.6 to 11.6) | 2.2 (0.4 to 4.7) |

Development values are out of sample. Deployment means *p<sub>s</sub>* ≥ 0.5, the net-benefit threshold. Parentheses give 95% bootstrap intervals; other sensitivities and false alarms are at development-set thresholds.

### 3.4. Personalisation from the index seizure

At the thresholds set on development patients, refitting increased pooled development sensi-tivity from 21.5% to 30.1%, while false alarms decreased from 2.0 to 1.5/day. In held-out pa-tients, sensitivity increased from 13.8% to 19.1% and false alarms from 0.39 to 0.77/day. No patient lost sensitivity, as shown in Fig. 4a–b, whereas false alarms rose in 29 of 70 develop-ment and 13 of 27 held-out patients.

**Fig. 4.**
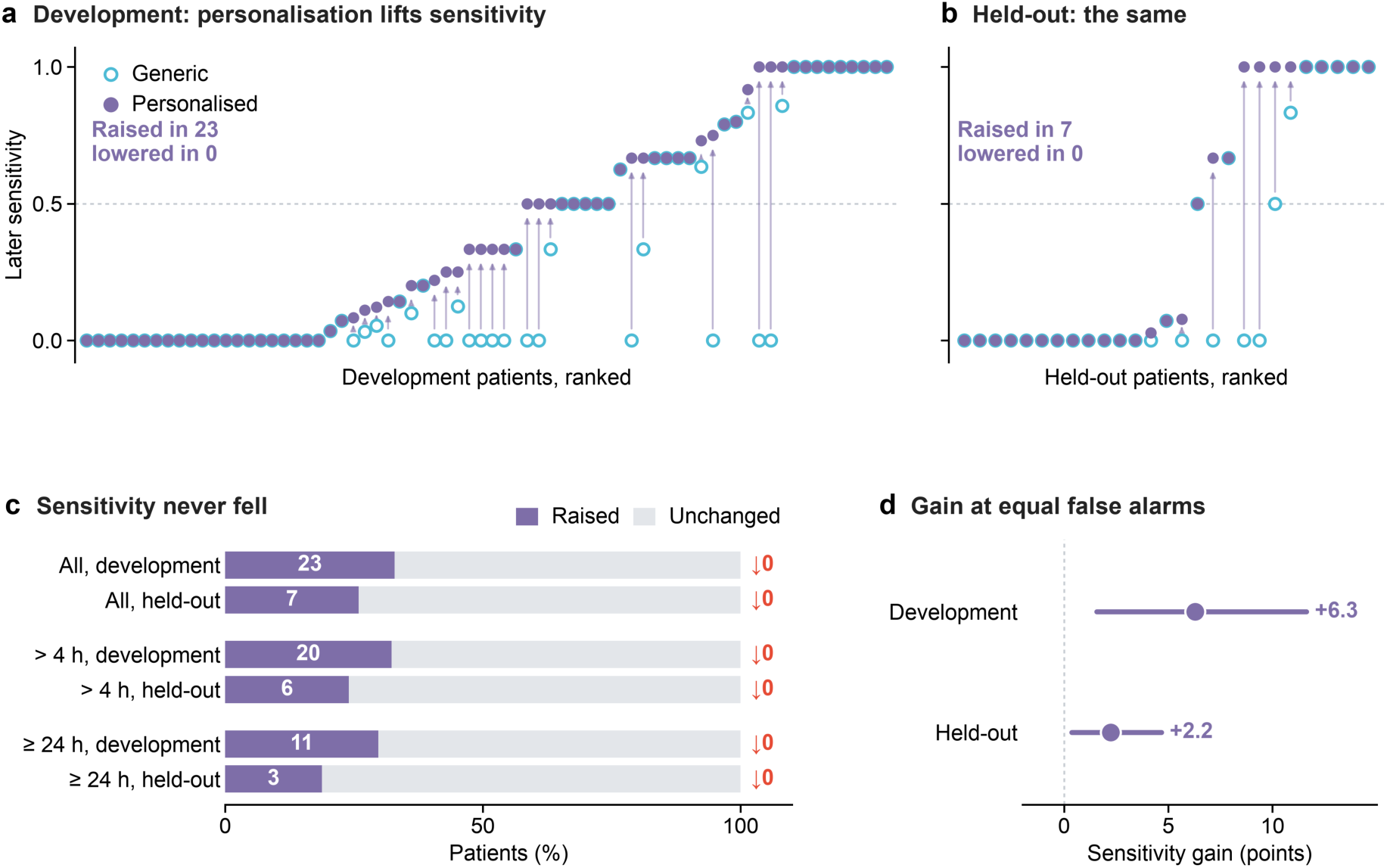
Personalisation improves later sensitivity. (a, b) Generic (open) and personalised (filled) sensitivity at de-velopment-set thresholds; arrows join increases and the dashed line marks 0.5. (c) Patient proportions with in-creased, unchanged or decreased sensitivity under each temporal restriction; numbers give increases and decreases. (d) Personalised minus generic sensitivity at equal false-alarm rates, averaged over 0.25–2/day, with 95% bootstrap intervals.

At equal false-alarm rates, averaged over 0.25–2 per day, personalisation raised later sensi-tivity by 6.3 percentage points in development patients, with a 95% confidence interval of 1.6–11.6. In held-out patients the gain was 2.2 points, with an interval of 0.4–4.7, as shown in Fig. 4d. Background or index-seizure windows alone gave smaller gains, as reported in Supplemen-tary Table S4. Beyond 4 h after the index seizure, sensitivity increased in six held-out patients and decreased in none, and beyond 24 h in three, as summarised in Fig. 4c.

## 4. Discussion

In this study, we used one recorded seizure for two connected decisions: predicting later sei-zure sensitivity and adapting the detector before deployment. Held-out qualification achieved AUC 0.74, and across all evaluable patients ProbeFit separated good-detection patients better than the heart-rate rule. At equal false-alarm rates, refitting raised later sensitivity in both sets.

This decision matters because users prioritise sensitivity and few false alarms, and uncertainty about accuracy affects continued wearable use [37, 38]. Sustained use has been reported for validated home detectors with low false-alarm rates [39]. ProbeFit supplies this information while video-EEG still provides a reference seizure.

Earlier heart-rate-based selection achieved high sensitivity in selected hospital patients [40], and responder status also distinguished performance in ambulatory ECG [41]. ProbeFit extends this approach from the seizure’s physiology to the detectors’ response. A multimodal response also carries the EEG information available to the model, and across all evaluable patients the probe outperformed the duration and type of the index seizure, which clinicians already read at the bedside.

Personalisation complements selection. Patient-specific wrist-sensor models [42] and intra-cranial EEG methods trained with one or a few seizures [43] demonstrate the value of individual seizure signatures. Transfer learning [44], few-sample meta-learning [45] and personalised fed-erated learning [46] offer further ways to reduce adaptation data. ProbeFit fixes when those data become available: refitting uses the index seizure and preceding background, and evaluation uses later seizures. The gain required both inputs, as neither alone matched their combination.

Seizure pathways vary over time, with similar pathways occurring closer together [47]. Nev-ertheless, discrimination persisted for seizures hours or a day after the index event, whereas the seizure-free control gave none. Qualification therefore reflects the detector’s ictal response ra-ther than the patient’s background score level or adjacent seizures.

The present evidence comes from inpatient recordings on one device. Medication withdrawal during monitoring changes seizure expression [48], while ambulatory recordings introduce movement artefacts and data loss [49]. Validation in longer home recordings is the next step under phased device-evaluation standards [50]. ProbeFit’s reach grows with the seizure types its detector covers, and short and non-motor seizures remain the main target. Demographic and centre-specific analyses require metadata absent from the public release, and a larger cohort will support recalibration and test whether the sensitivity-based selection decision translates into sustained home use.

## 5. Conclusions

ProbeFit uses the first recorded seizure to combine patient selection and detector personali-sation during the first monitoring admission. Its qualification probability predicted later seizure sensitivity in held-out patients, and refitting with the same seizure and its preceding background raised sensitivity at equal false-alarm rates.

## Ethics statement

This study analysed the public, de-identified SeizeIT2 dataset. The original data collection was approved by the ethics committees of the participating centres and all participants gave in-formed consent; no new data were collected, so no further approval was required.

## CRediT authorship contribution statement

**Han Cui:** Conceptualization, Methodology, Software, Formal analysis, Visualization, Writing – original draft. **Yuge Zhu:** Validation, Writing – review & editing. **Shaojia Huang:** Supervi-sion, Writing – review & editing. **Tao Xu:** Supervision, Project administration, Funding acqui-sition, Writing – review & editing.

## Data and code availability

The SeizeIT2 data are publicly available on OpenNeuro (ds005873). The code for all analyses and figures will be made publicly available upon journal publication.

## Declaration of generative AI and AI-assisted technologies in the manuscript preparation process

During the preparation of this work, the authors used Claude (Anthropic) to assist in reviewing code and improving readability. After using this tool, the authors reviewed and edited the con-tent as needed and take full responsibility for the content of the publication.

## Data Availability

The SeizeIT2 data analysed in this study are publicly available on OpenNeuro (ds005873, https://openneuro.org/datasets/ds005873). The code for all analyses and figures will be made publicly available upon journal publication.

## References

[1] Beghi E, Giussani G, Nichols E, Abd-Allah F, Abdela J, Abdelalim A, et al. Global, regional, and national burden of epilepsy, 1990-2016: a systematic analysis for the Global Burden of Disease Study 2016. Lancet Neurol 2019;18:357–75.

[2] Fisher RS, Acevedo C, Arzimanoglou A, Bogacz A, Cross JH, Elger CE, et al. ILAE official report: a practi-cal clinical definition of epilepsy. Epilepsia 2014;55:475–82.

[3] Fisher RS, Cross JH, French JA, Higurashi N, Hirsch E, Jansen FE, et al. Operational classification of seizure types by the International League Against Epilepsy: position paper of the ILAE Commission for Classifica-tion and Terminology. Epilepsia 2017;58:522–30.

[4] Beniczky S, Wiebe S, Jeppesen J, Tatum WO, Brazdil M, Wang Y, et al. Automated seizure detection using wearable devices: a clinical practice guideline of the International League Against Epilepsy and the Interna-tional Federation of Clinical Neurophysiology. Epilepsia 2021;62:632–46.

[5] Bernini A, Dan J, Ryvlin P. Ambulatory seizure detection. Curr Opin Neurol 2024;37:99–104.

[6] Macea J, Bhagubai M, Broux V, De Vos M, Van Paesschen W. In-hospital and home-based long-term moni-toring of focal epilepsy with a wearable electroencephalographic device: diagnostic yield and user experi-ence. Epilepsia 2023;64:937–50.

[7] Hoppe C, Poepel A, Elger CE. Epilepsy: accuracy of patient seizure counts. Arch Neurol 2007;64:1595–9.

[8] Elger CE, Hoppe C. Diagnostic challenges in epilepsy: seizure under-reporting and seizure detection. Lancet Neurol 2018;17:279–88.

[9] Ryvlin P, Nashef L, Lhatoo SD, Bateman LM, Bird J, Bleasel A, et al. Incidence and mechanisms of cardi-orespiratory arrests in epilepsy monitoring units (MORTEMUS): a retrospective study. Lancet Neurol 2013;12:966–77.

[10] Dong C, van Dijk JP, Aarts RM, Wang Y, Long X. Detection of nocturnal epileptic seizures using a weara-ble armband: a deep learning approach combining accelerometry and photoplethysmography signals. Com-put Methods Programs Biomed 2026;273:109087.

[11] Vandecasteele K, De Cooman T, Dan J, Cleeren E, Van Huffel S, Hunyadi B, et al. Visual seizure annota-tion and automated seizure detection using behind-the-ear electroencephalographic channels. Epilepsia 2020;61:766–75.

[12] Vandecasteele K, De Cooman T, Chatzichristos C, Cleeren E, Swinnen L, Macea Ortiz J, et al. The power of ECG in multimodal patient-specific seizure monitoring: added value to an EEG-based detector using limited channels. Epilepsia 2021;62:2333–43.

[13] Zijlmans M, Flanagan D, Gotman J. Heart rate changes and ECG abnormalities during epileptic seizures: prevalence and definition of an objective clinical sign. Epilepsia 2002;43:847–54.

[14] Eggleston KS, Olin BD, Fisher RS. Ictal tachycardia: the head-heart connection. Seizure 2014;23:496–505.

[15] Bhagubai M, Chatzichristos C, Swinnen L, Macea J, Zhang J, Lagae L, et al. SeizeIT2: wearable dataset of patients with focal epilepsy. Sci Data 2025;12:1228.

[16] Wu K, Zhao Z, Yener B. Large EEG-U-Transformer for time-step level detection without pre-training. arXiv:2504.00336v3 [Preprint] 2025.

[17] You S, Cho BH, Yook S, Kim JY, Shon YM, Seo DW, et al. Unsupervised automatic seizure detection for focal-onset seizures recorded with behind-the-ear EEG using an anomaly-detecting generative adversarial network. Comput Methods Programs Biomed 2020;193:105472.

[18] Dan J, Pale U, Amirshahi A, Cappelletti W, Ingolfsson TM, Wang X, et al. SzCORE: seizure community open-source research evaluation framework for the validation of electroencephalography-based automated seizure detection algorithms. Epilepsia 2025;66:14–24.

[19] Qu H, Gotman J. A patient-specific algorithm for the detection of seizure onset in long-term EEG monitoring: possible use as a warning device. IEEE Trans Biomed Eng 1997;44:115–22.

[20] Shoeb A, Edwards H, Connolly J, Bourgeois B, Treves ST, Guttag J. Patient-specific seizure onset detection. Epilepsy Behav 2004;5:483–98.

[21] Giannakakis G, Tsiknakis M, Vorgia P. Focal epileptic seizures anticipation based on patterns of heart rate variability parameters. Comput Methods Programs Biomed 2019;178:123–33.

[22] Sopic D, Teijeiro T, Atienza D, Aminifar A, Ryvlin P. Personalized seizure signature: an interpretable ap-proach to false alarm reduction for long-term epileptic seizure detection. Epilepsia 2023;64 Suppl 4:S23–33.

[23] Jeppesen J, Christensen J, Johansen P, Beniczky S. Personalized seizure detection using logistic regression machine learning based on wearable ECG-monitoring device. Seizure 2023;107:155–61.

[24] You S, Cho BH, Shon YM, Seo DW, Kim IY. Semi-supervised automatic seizure detection using personal-ized anomaly detecting variational autoencoder with behind-the-ear EEG. Comput Methods Programs Bio-med 2022;213:106542.

[25] Jeppesen J, Fuglsang-Frederiksen A, Johansen P, Christensen J, Wüstenhagen S, Tankisi H, et al. Seizure detection based on heart rate variability using a wearable electrocardiography device. Epilepsia 2019;60:2105–13.

[26] Jeppesen J, Fuglsang-Frederiksen A, Johansen P, Christensen J, Wüstenhagen S, Tankisi H, et al. Seizure detection using heart rate variability: a prospective validation study. Epilepsia 2020;61 Suppl 1:S41–6.

[27] Jeppesen J, Christensen J, Ahrenfeldt Petersen O, Fenger S, Armand Larsen S, Wüstenhagen S, et al. Sei-zure detection using wearable electrocardiogram connected to a smartphone: a phase 3 clinical validation study. eBioMedicine 2025;120:105952.

[28] Reintjes C, Hagenbeck JF, Ballo M, Rahlmeier T, Wolf SM, Schoder D. ECG-based detection of epileptic seizures in real-world wearable settings: insights from the SeizeIT2 dataset. Sensors 2025;25:7687.

[29] Alhaskir M, Kutafina E, Linke F, Fischer FP, Schriewer E, Lauxmann S, et al. Reliable detection of focal onset impaired awareness seizures in patients with epilepsy using wearable ECG: development and valida-tion study. Comput Methods Programs Biomed 2026;279:109295.

[30] Swinnen L, Bhagubai M, Chatzichristos C, et al. A multicenter, video-EEG-based validation of a multi-modal wearable device for focal seizure detection in adults: the SeizeIT2 study. Epilepsia Open 2026;11:883–94.

[31] Seoni S, Molinari F, Benevieri M, Salvi M. An integrated calibration–uncertainty framework for improving the reliability of deep learning models for seizure detection in EEG signals. Comput Methods Programs Bi-omed 2026;285:109520.

[32] Friedman JH. Greedy function approximation: a gradient boosting machine. Ann Stat 2001;29:1189–232.

[33] Pan J, Tompkins WJ. A real-time QRS detection algorithm. IEEE Trans Biomed Eng 1985;32:230–6.

[34] Vickers AJ, Elkin EB. Decision curve analysis: a novel method for evaluating prediction models. Med Decis Making 2006;26:565–74.

[35] Van Calster B, McLernon DJ, van Smeden M, Wynants L, Steyerberg EW. Calibration: the Achilles heel of predictive analytics. BMC Med 2019;17:230.

[36] Collins GS, Moons KGM, Dhiman P, Riley RD, Beam AL, Van Calster B, et al. TRIPOD+AI statement: updated guidance for reporting clinical prediction models that use regression or machine learning methods. BMJ 2024;385:e078378.

[37] Bruno E, Simblett S, Lang A, Biondi A, Odoi C, Schulze-Bonhage A, et al. Wearable technology in epi-lepsy: the views of patients, caregivers, and healthcare professionals. Epilepsy Behav 2018;85:141–9.

[38] Bruno E, Viana PF, Sperling MR, Richardson MP. Seizure detection at home: do devices on the market match the needs of people living with epilepsy and their caregivers? Epilepsia 2020;61 Suppl 1:S11–24.

[39] Hadady L, Klivényi P, Fabó D, Beniczky S. Real-world user experience with seizure detection wearable devices in the home environment. Epilepsia 2023;64 Suppl 4:S72–7.

[40] Jeppesen J, Lin K, Melo HM, Pavei J, Marques JLB, Beniczky S, et al. Detection of seizures with ictal tach-ycardia, using heart rate variability and patient adaptive logistic regression machine learning methods: a hospital-based validation study. Epileptic Disord 2024;26:199–208.

[41] Li J, Nurse ES, Grayden DB, Cook MJ, Karoly PJ. Epileptic seizure detection using heart rate variability from ambulatory ECG: a pseudoprospective study. J Neural Eng 2025;22:026033.

[42] Böttcher S, Bruno E, Epitashvili N, Dümpelmann M, Zabler N, Glasstetter M, et al. Intra- and inter-subject perspectives on the detection of focal onset motor seizures in epilepsy patients. Sensors (Basel) 2022;22:3318.

[43] Burrello A, Schindler K, Benini L, Rahimi A. Hyperdimensional computing with local binary patterns: one-shot learning of seizure onset and identification of ictogenic brain regions using short-time iEEG record-ings. IEEE Trans Biomed Eng 2020;67:601–13.

[44] Nasseri M, Pal Attia T, Joseph B, Gregg NM, Nurse ES, Viana PF, et al. Non-invasive wearable seizure de-tection using long-short-term memory networks with transfer learning. J Neural Eng 2021;18:056017.

[45] Amirshahi A, Toosi MH, Mohammadi S, Albini S, Schiavone PD, Ansaloni G, et al. A meta-learning method with reduced data requirements for training and updating deep learning models on wearable de-vices. Commun Med (Lond) 2026;6:523.

[46] Baghersalimi S, Teijeiro T, Atienza D, Aminifar A. Personalized real-time federated learning for epileptic seizure detection. IEEE J Biomed Health Inform 2022;26:898–909.

[47] Schroeder GM, Diehl B, Chowdhury FA, Duncan JS, de Tisi J, Trevelyan AJ, et al. Seizure pathways change on circadian and slower timescales in individual patients with focal epilepsy. Proc Natl Acad Sci U S A 2020;117:11048–58.

[48] Duy PQ, Krauss GL, Crone NE, Ma M, Johnson EL. Antiepileptic drug withdrawal and seizure severity in the epilepsy monitoring unit. Epilepsy Behav 2020;109:107128.

[49] Böttcher S, Vieluf S, Bruno E, Joseph B, Epitashvili N, Biondi A, et al. Data quality evaluation in wearable monitoring. Sci Rep 2022;12:21412.

[50] Beniczky S, Ryvlin P. Standards for testing and clinical validation of seizure detection devices. Epilepsia 2018;59 Suppl 1:9–13.

