## supplementary_information for "ProbeFit: one-seizure qualification and personalisation of multimodal wearable seizure detectors"

### Supplementary material

#### S1. Detector preprocessing and training

The gradient-boosted detector works on hand-crafted features of 2-s windows. For each bte-EEG channel we computed log power in six bands between 1 and 95 Hz, relative power in the first five, line length, spectral peak ratio, spectral entropy and log variance. From the ECG we derived heart rate, its difference from the median of the preceding 5 min, its 30-s change and its 10-s variability. The EMG gave log root-mean-square amplitude and high-frequency power, and the movement sensors the standard deviation of acceleration and rotation. Each window also received the mean of its 5-window neighbourhood and its deviation from the 30-window mean, so the classifier sees the local temporal context. A histogram-based gradient-boosting classifier with 300 iterations, learning rate 0.08 and 31 leaves per tree was trained on all seizure windows and a random 2% of non-seizure windows, with class weights balancing the two.

SeizureTransformer is a published convolutional and transformer network that labels every time step of a 60-s window. We retrained it from random initialisation on three channels, the two EEG channels and the ECG, at 256 Hz after 0.5–120 Hz band-pass filtering, 50-Hz notch filtering and robust scaling by the median and median absolute deviation of each recording run. Training used binary cross-entropy, the RAdam optimiser with learning rate $10^{-4}$, and early stopping on the window-level precision-recall area of 10% of the training patients. Per-sample outputs were averaged to 2-s windows.

#### S2. PROBEFIT ALGORITHM

**Algorithm S1. ProbeFit.** Input: the recording $X_0$ covering the index seizure and its scoring margin, the generic detectors G and T, and the training sample D of other patients. Output: a deployment decision and the index-personalised detector R.

Step 1: score the 2-s windows of $X_0$ with G and T and smooth the scores with a centred five-window mean.

Step 2: take each detector's peak score $m_k$ from 30 s before the onset to 60 s after the end of the index seizure.

Step 3: clip the peaks, take logits by Eq. (2), standardise them and combine them into $p_s$ by Eq. (3).

Step 4: refit G on D, the index-seizure windows and up to 20,000 pre-index background windows, weighted as in Eq. (4), to obtain R.

Step 5: if $p_s$ passes the gate of Eq. (5), deploy R with threshold $\tau_R$; otherwise wait.

#### SUPPLEMENTARY TABLES

**Table S1. Detection of later seizures by seizure type, duration and vigilance.**

| Later seizures | Development n | Generic (%) | Personalised (%) | Held-out n | Generic (%) | Personalised (%) |
|---|---|---|---|---|---|---|
| Focal to bilateral tonic-clonic | 16 | 94 | 94 | 17 | 82 | 88 |
| Focal motor | 283 | 25 | 33 | 37 | 16 | 24 |
| Focal non-motor | 219 | 11 | 21 | 68 | 1 | 7 |
| Focal, motor signs not specified | 86 | 23 | 34 | 25 | 0 | 0 |
| Unknown onset | 1 | 0 | 0 | 5 | 0 | 0 |
| Duration ≤ 20 s | 252 | 2 | 7 | 67 | 1 | 3 |
| Duration > 20 s | 353 | 35 | 46 | 85 | 24 | 32 |
| During sleep | 254 | 24 | 28 | 84 | 10 | 13 |
| Awake | 282 | 21 | 32 | 64 | 16 | 23 |
| All | 605 | 21 | 30 | 152 | 14 | 19 |

Detection at the thresholds set on development patients. Types follow the release annotations; focal motor excludes focal to bilateral tonic-clonic seizures. Vigilance was not annotated for every seizure.

**Table S2. Discrimination of good-detection patients by ProbeFit and by predictors read from the index seizure.**

| Predictor | Development AUC | Held-out AUC | All 97 patients AUC |
|---|---|---|---|
| ProbeFit $p_s$ | 0.77 (0.66 to 0.88) | 0.74 (0.52 to 0.91) | 0.76 (0.66 to 0.85) |
| Gradient-boosted peak | 0.73 (0.60 to 0.84) | 0.75 (0.53 to 0.93) | 0.73 (0.62 to 0.82) |
| SeizureTransformer peak | 0.80 (0.69 to 0.90) | 0.74 (0.54 to 0.92) | 0.78 (0.68 to 0.87) |
| Heart-rate rule | 0.59 (0.47 to 0.70) | 0.59 (0.41 to 0.78) | 0.59 (0.49 to 0.69) |
| Index seizure detected by the generic detector | 0.60 (0.51 to 0.70) | 0.68 (0.51 to 0.83) | 0.62 (0.54 to 0.70) |
| Index-seizure duration | 0.66 (0.53 to 0.79) | 0.75 (0.55 to 0.92) | 0.69 (0.58 to 0.79) |
| Index-seizure duration and type | 0.56 (0.42 to 0.69) | 0.82 (0.63 to 0.97) | 0.64 (0.53 to 0.75) |
| **Net benefit at threshold 0.5** | | | |
| Deployment by $p_s \geq 0.5$ | 0.16 (0.00 to 0.31) | 0.07 (−0.22 to 0.33) | 0.13 (0.00 to 0.27) |
| Heart-rate rule | 0.04 (−0.11 to 0.21) | 0.04 (−0.22 to 0.30) | 0.04 (−0.09 to 0.19) |
| Index seizure detected | 0.09 (−0.01 to 0.19) | 0.15 (0.00 to 0.33) | 0.10 (0.02 to 0.20) |
| Duration and type, probability ≥ 0.5 | 0.01 (−0.11 to 0.14) | 0.19 (−0.04 to 0.44) | 0.06 (−0.06 to 0.19) |
| Deployment to all | −0.09 (−0.31 to 0.17) | −0.11 (−0.48 to 0.26) | −0.09 (−0.30 to 0.11) |

Values are estimates with 95% patient-level bootstrap intervals. Development probabilities are leave-one-patient-out; the duration-and-type model (log duration, motor signs, bilateral tonic-clonic evolution and impaired awareness) was fitted in the same way and applied unchanged to held-out patients. In all 97 patients, adding the two detector peaks to duration and type improved fit (likelihood-ratio $\chi^2 = 13.8$, 2 df, $p = 0.001$), whereas adding duration and type to the peaks did not ($\chi^2 = 3.1$, 4 df, $p = 0.55$).

**Table S3. Robustness of qualification to the outcome definition.**

| Analysis | Development | Held-out | All 97 patients |
|---|---|---|---|
| AUC, later sensitivity ≥ 1/3 | 0.73 | 0.74 | 0.72 |
| AUC, later sensitivity ≥ 1/2 | 0.77 | 0.74 | 0.76 |
| AUC, later sensitivity ≥ 2/3 | 0.81 | 0.71 | 0.78 |
| AUC, later sensitivity ≥ 3/4 | 0.80 | 0.70 | 0.77 |
| AUC, patients with ≥ 2 later seizures | 0.73 (0.59 to 0.86), n = 55 | 0.85 (0.60 to 1.00), n = 16 | 0.75 (0.64 to 0.86), n = 71 |
| Later seizures detected, deployed / not deployed (%) | 51 / 14 | 26 / 12 | 46 / 14 |

Seizure-level rates pool all later seizures of the patients in each group, at the personalised detector's threshold.

**Table S4. Sensitivity gain over the generic detector at equal false-alarm rates (percentage points).**

| Refitting data | Development, 1/day | Development, 0.25–2/day | Held-out, 1/day | Held-out, 0.25–2/day |
|---|---|---|---|---|
| Index seizure and background (ProbeFit) | 6.9 (0.2 to 12.8) | 6.3 (1.6 to 11.6) | 0.0 (−1.9 to 4.5) | 2.2 (0.4 to 4.7) |
| Background only | 4.5 (−0.5 to 9.1) | 4.6 (0.5 to 9.6) | −4.0 (−6.6 to 0.0) | −1.3 (−2.9 to −0.1) |
| Index seizure only | −10.9 (−21.9 to 0.9) | −10.6 (−20.3 to 0.7) | −1.3 (−3.2 to 2.5) | 1.0 (−0.5 to 3.3) |

For each detector, the threshold was the lowest value giving the target pooled false-alarm rate. Intervals come from patient-level bootstrap resamples in which thresholds were reset (5,000 resamples at 1/day; 1,000 for the average over 0.25–2/day).
